# Early-life exposure to perfluorinated alkyl substances modulates lipid metabolism in progression to celiac disease

**DOI:** 10.1101/2020.04.02.20051359

**Authors:** Lisanna Sinisalu, Partho Sen, Samira Salihović, Suvi M. Virtanen, Heikki Hyöty, Jorma Ilonen, Jorma Toppari, Riitta Veijola, Matej Orešič, Mikael Knip, Tuulia Hyötyläinen

## Abstract

**OBJECTIVES:** Celiac disease (CD) is a systemic immune-mediated disorder with increased frequency in the developed countries over the last decades implicating the potential causal role of various environmental triggers in addition to gluten. Herein, we apply determination of perfluorinated alkyl substances (PFAS) and combine the results with the determination of bile acids (BAs) and molecular lipids, with the aim to elucidate the impact of prenatal exposure on risk of progression to CD in a prospective series of children prior the first exposure to gluten (at birth and at 3 months of age).

**METHODS:** We analyzed PFAS, BAs and lipidomic profiles in 76 plasma samples at birth and at 3 months of age in the Type 1 Diabetes Prediction and Prevention (DIPP) study (n=17 progressors to CD, n=16 healthy controls, HCs).

**RESULTS:** Plasma PFAS levels showed a significant inverse association with the age of CD diagnosis in infants who later progressed to the disease. Associations between BAs and triacylglycerols (TGs) showed different patterns already at birth in CD progressors, indicative of different absorption of lipids in these infants.

**DISCUSSION:** PFAS exposure may modulate lipid and BA metabolism, and the impact is different in the infants who develop CD later in life, in comparison to HCs. The results indicate more efficient uptake of PFAS in such infants. Higher PFAS exposure during prenatal and early life may accelerate the progression to CD in the genetically predisposed children.

**Study Highlights:** *WHAT IS KNOWN:* Several observational studies have implicated a role of early life environmental triggers other than gluten in the development of CD. This is supported by the findings showing dysregulation of lipids already prior to the first introduction of gluten.

*WHAT IS NEW HERE:* We show that prenatal exposure to perfluorinated compounds is associated with changes in the lipid metabolism, most likely through the bile acids, and that a high exposure during prenatal and early life may accelerate the progression to CD in the genetically predisposed children.

*TRANSLATIONAL IMPACT:* Exposure to environmental chemicals may impact the rate of progression to CD and should be assessed as a potential risk factor of CD in larger clinical cohort settings.

## Introduction

Celiac disease (CD) is a systemic immune-mediated disorder, which is triggered by gluten and other prolamins in genetically susceptible individuals^1^. The frequency of CD has increased in the developed countries over the last decades, implicating the potential causal role of various environmental triggers in addition to gluten. Similarly to CD, the rate of other autoimmune diseases such as type 1 diabetes (T1D) has also increased during the last decades^2,3^. CD and T1D share common predisposing alleles in the class II HLA-region^4,5^, and approximately 10% of patients with T1D also develop clinical CD^6,7^, while subjects with CD are at-risk for developing T1D before 20 years of age^8^. The possible triggers that have been indicated to affect the progression to CD include the composition of the intestinal microbiota, infant feeding, and the use of antibiotics^1,9,10^.

The hygiene hypothesis, stating that a decrease of the infectious burden is associated with the rise of allergic and autoimmune diseases, has also been proposed in CD because CD has shown to be more common in developed countries^11-13^. Another possible explanation for varying incidence in different populations implicates the role of different infant feeding patterns (including the amount and timing of gluten introduction) in families with low socioeconomic status^12^. Socioeconomical status (SES) may also have a broader role in increasing the risk for CD, such as different exposures to environmental pollutants. Higher SOS has in several studies been linked to higher burdens of several persistent organic pollutants (POP), particularly to perfluorinated alkyl substances (PFAS)^14,15^. Exposure to POPs has, however, been poorly investigated as a risk factor for CD. In T1D, recent studies identified associations between the exposures to environmental chemicals and the increased risk of islet autoimmunity^16-19^. We have shown that fetal exposure to PFAS increases the risk of T1D in genetically susceptible individuals^20^, and further, that the metabolic profiles of the infants are affected by the prenatal exposure to PFAS. In particular, the PFAS exposure has a marked impact on bile acid (BA) profiles, which, in turn, were linked to changes in specific phospholipids, *e*.*g*., lysophosphatodylcholines (LPCs), phosphatodylcholines (PCs) and sphingomyelins (SMs). Similarly, we have recently identified systematic differences in plasma lipid profiles between children who later progressed to clinical CD during the follow-up, when compared to children who remained healthy^5^. Importantly, these differences were observed prior to the first introduction of gluten in the diet and before the first signs of CD-associated autoimmunity. Similar results were recently reported in an Italian cohort study^21^.

In line with the observations of intestinal dysbiosis in CD^1^, changes in lipid metabolism^5,21^ as well as the documented link of CD with liver disorders^22^, it has been shown that circulating BAs are elevated in CD, including in children^23,24^. BAs not only facilitate the digestion and absorption of dietary lipids in the small intestine, they are also important metabolic regulators involved in the maintenance of lipid and glucose homeostasis^25^. BAs are produced in the liver, and their homeostasis is maintained through the tightly controlled enterohepatic circulation. Moreover, there is a close interplay between BA and gut microbiota. Gut microbiota is involved in the biotransformation of secondary BAs, while BAs can modulate the microbial composition in the gut due to their antimicrobial activity as well as through regulation of the composition of the intestinal microbiota through farnesoid X receptor (FXR) and Takeda G-protein receptor 5 (TGR-5)^26,27^. PFAS, on the other hand, have been shown to undergo similar enterohepatic circulation as the BAs, and PFAS can also suppress the BA biosynthesis in the liver^28^.

Here we hypothesized that early life PFAS exposure affects the BA profiles in children and explains the association between the early changes in lipid profiles in those children who later progress to clinical CD. We investigated the impact of early-life exposure to PFAS in children who progressed to clinical CD during their follow-up, and compared the exposures to those observed in children who remained healthy during the follow-up. In addition, we also analyzed BAs in the same samples, and performed integrative analysis of previously published lipidomic profiles^5^, together with PFAS and BA levels.

## Methods

### Study design

The current study is a part of the Type 1 Diabetes Prediction and Prevention study in Finland (DIPP), which is an ongoing prospective birth cohort study initiated in 1994. In DIPP, parents of newborn children at the university hospitals of Turku, Tampere and Oulu in Finland are asked for permission to screening for T1D-conferring HLA risk alleles in the umbilical cord blood. Families of children identified as having an increased HLA-conferred risk for T1D were invited to join the follow-up study. This study included children born in Tampere University Hospital between August 1999 and September 2005. During that period, 23,839 children were screened at birth for increased risk of T1D, and 2,642 eligible children were enrolled in the follow-up and had at least two visits to the study center. These children carry the high-risk HLA DQB1*02/*03:02 genotype or the moderate-risk HLA-DQB1*03:02/x genotype (x ≠ DQB1*02, 03:01, or 06:02). More than 1,200 of these children took part in the DIPP-CD study. Because of the enrolment criteria 60% of the children were male. At each visit, the families were interviewed for dietary changes, infections, growth, important family related issues and the children gave a non-fasting venous blood sample. The children were followed for four CD-related antibodies: anti-transglutaminase 2 (anti-TG2), anti-endomysium (EMA), antigliadin (AgA-IgG and AgA-IgA) and anti-reticulin (ARA) antibodies, and for four T1D-associated autoantibodies: islet cell autoantibodies (ICA), autoantibodies against insulin (IAA), tyrosine phosphatase-like protein (IA-2A) and glutamate decarboxylase (GADA). IgA deficiency was excluded. All children participating in this study were of Caucasian origin. None of the mothers had CD. The children in the CD follow-up cohort were annually screened for anti-TG2 antibodies using a commercial kit (Celikey Pharmacia Diagnostics, Freiburg, Germany). If a child’s sample was found positive, all previous and forthcoming samples were analyzed for the entire set of CD-related antibodies. A duodenal biopsy was recommended for all anti-TG2-positive children. If the biopsy was consistent with the ESPGHAN criteria of 1990, a gluten-free diet (GFD) was recommended. None of the children participating in this study had any T1D-associated autoantibodies in any samples during the follow-up. The first exposure to gluten in this study was at the median age of 5 months (**Table 1**).

**Table 1.** Demographic and clinical characteristics of the study subjects.

|  | **Celiac Disease** | **Healthy Controls** |
| --- | --- | --- |
| **Number of subjects** | 17 | 16 |
| **Gender** |  |  |
| Male (median) | 10 | 8 |
| Female (median) | 7 | 8 |
| **Age of first gluten intake**<br>(median) | 5.25 months | 5.3 months |
| **Age of first tTGA**<br>(median)(seroconversion) | 58 months | -- |
| **First tTGA level in plasma**<br>(median EIU) | 116.8 | -- |
| **Endoscopy Age**<br>(median) | 58.5 months | -- |
| **tTGA level after GFD**<br>(median EIU) | 7.1 | -- |
| **Total follow-up age from birth**<br>(median) | 6.08 years | 6.2 years |

We have randomly selected 17 children with biopsy proven CD (progressors) and a matched control for each progressor, carrying similar risk HLA alleles, born within ±1 month of each other, having had each sample taken within ±1 month of each other and living in the same region of the country throughout the whole follow-up period. Samples at birth (cord blood) and at 3 months of age were analyzed. Altogether, 76 plasma samples from children developing CD and from their HCs were analyzed. The clinical and genetic data of the participants are found in **Table 1**.

The ethics committee of the Tampere University Hospital approved the study. Written informed consent was obtained from the parents for HLA-screening, autoantibody analysis and intestinal biopsies.

### Analysis of PFAS and BAs

The BAs and PFAS were analyzed using the established method as described previously^29^. In brief, the sample preparation was done with 25 mg Ostro Protein Precipitation and Phospholipid Removal 96-well plate (Waters Corporation, Milford, USA), using 30 μL of serum and a set of BA and PFAS internal standards. Matrix-matched calibration standards were made using newborn bovine serum. Analyses were performed on an Acquity UPLC system coupled to a triple quadrupole mass spectrometer (Waters Corporation, Milford, USA) with an atmospheric electrospray interface operating in negative ion mode. Aliquots of 10 μL of samples were injected into the Acquity UPLC BEH C18 2.1 mm × 100 mm, 1.7 μm column (Waters Corporation). A trap column (PFC Isolator column, Waters Corporation) was installed between the pump and injector and used to retain fluorinated compounds originating from the HPLC system and the mobile phase. The eluent system consisted of (A) 2 mM NH4Ac in 70% MiliQ: 30% methanol and (B) 2 mM NH4Ac in 100% methanol. The gradient was programmed as follows: 0–1 min, 1 % solvent B; 1–13 min, 100 % solvent B; 13-16 min, 100 % solvent B; 16-17 min, 1 % solvent B, flow rate 0.3 mL/min. The total run time for UPLC-MS/MS analysis was 17 minutes, while the total run time for each sample injection was 20 minutes, including the reconditioning of the analytical column. MS analysis was performed in multiple reaction monitoring (MRM) mode and experimental details of the MS/MS method are given in **Supplementary Table S1**.

### Lipidomic analyses

The lipidomic analyses were performed as described previously^5^. The plasma samples (10 μL) were extracted using a modified version of the previously published Folch procedure^30^. The samples were analyzed using an ultra-high-performance liquid chromatography quadrupole time-of-flight mass spectrometry method (UHPLC-Q-TOF-MS from Agilent Technologies (Santa Clara, CA, USA). The analysis was carried out on an ACQUITY UPLC® BEH C18 column (2.1 mm × 100 mm, particle size 1.7 μm) by Waters (Milford, USA). Internal standard mixture was used for normalization and lipid-class specific calibration was used for quantitation as previously described. Quality control was performed throughout the dataset by including blanks, pure standard samples, extracted standard samples and control plasma samples. Relative standard deviations (%RSDs) for lipid standards representing each lipid class in the control plasma samples (*n*=8) and in the pooled serum samples (*n* = 20) were on average 11.7% (raw variation). The lipid concentrations in the pooled control samples was on average 8.4% and 11.4% in the standard samples. This shows that the method is reliable and repeatable throughout the sample set. MS data processing was performed using open source software MZmine 2.18^31^.

### Statistical analyses

Statistical analysis was performed using R (v3.6.0) statistical programming language^32^. Previously published lipidomics dataset^5^ was obtained from *MetaboLights* with the study identifier *MTBLS729*. The data was log-transformed and missing values were imputed by half of the row’s minimum. The datasets were auto-scaled prior to multivariate analyses.

To integrate across different data types, we applied sparse generalized correlation discriminant analysis *via* the DIABLO framework, part of *mixOmics* package (*v6*.*10*.*8*)^33^. The method constructs components across different data blocks, by maximizing their covariance with each other and a given response (Y) variable. Heterogenous data such as PFAS, BAs and lipid levels were partitioned into three different blocks and regressed to a binary response variable, *i*.*e*., CD progressors (CD) or HCs. Regularized sparse partial least squares discriminant (sPLS-DA)^33,34^ models were fitted. The optimal number of components that achieve the best performance based on the overall error rate or Balanced Error Rate (BER) were determined. Block sPLSDA models were developed at two time-points, *i*.*e*., at birth (cord blood) and at 3 months of age. Moreover, these models were cross-validated^35^ by 5-fold cross-validation (CV) with (N =100 repeats). The final model performances were assessed by area under the curve (AUC), overall misclassification error rate, and BER generated using *‘perf’,’auroc’,’block*.*splsda’* functions.

The key predictors/contributors that are jointly associated with the response variable of interest across all the input data matrices were identified by their *Variable Importance (VI)* score, i.e (absolute weighted loadings). The direction (up or down) of contribution of a particular predictor was determined by estimating the mean relative log fold change in the CD progressors vs. HCs at a particular time-point.

In order to understand which variables influence the outcomes, multivariate correlations were performed by a method described in^33^. In addition, bivariate correlation was performed using Pearson correlation. Debiased Sparse Partial Correlation algorithm (DSPC) was used for estimating partial correlation networks, visualized by the *MetaboAnalyst* 436 version with cutoff values of correlations between +/- 0.22 to 0.75. Univariate analysis (unpaired two sample t-test and paired t-test) using the *‘t*.*test’* function, was deployed to identify mean differences in the concentration of individual PFAS and BAs between CD progressors *vs*. HCs at birth and at 3 months of age. Libraries/packages such as *‘Heatmap*.*2’*, ‘*mixOmics’, ‘boxplot’, ‘beanplot’*, ‘*gplot*’ and ‘*ggplot2*’ were used for data visualization.

## Results

### Levels of PFAS and BAs in the infants

Seven PFAS compounds were detected in the samples, namely PFHpA, PFHxS, PFOA, PFNA, PFOS, PFDA and PFUnDA, both at birth and at 3 months of age (**Supplementary Table S2**). PFOS and PFOA had the highest concentrations and they were detected in the majority of samples, whilst PFDA and PFNA were detected in less than 10% of the samples and were excluded from further analyses. The levels of PFAS were lower in the cord blood (CB) than at the age of 3 months **(Figure 1A-C)**. No significant differences were observed between cases and controls either at birth or at 3 months of age **(Figure 1A-C)**. Interestingly, the levels of the total PFAS were significantly elevated from birth to 3 months of age only in the CD group (fold change (FC) 3 months vs. at birth = 1.72, p = 0.003 **Supplementary Table S3**) although also controls had an increasing trend (FC = 1.02, p = 0.35). Among the individual PFAS, PFOA showed nearly twofold increase, being significant in both groups **(Figure 1A-C)** (**Supplementary Table S3**).

**Figure 1.**
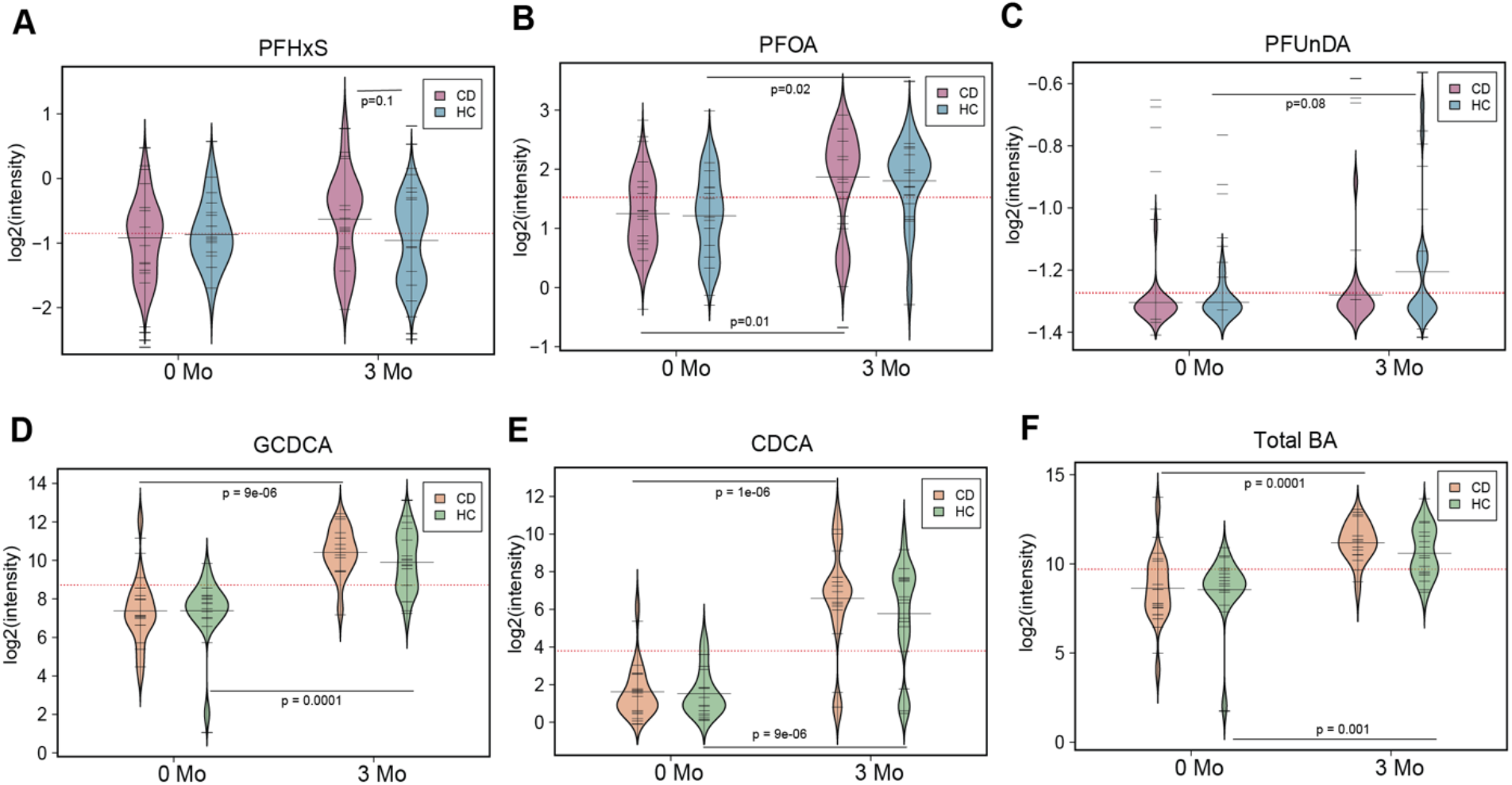
Bean plots showing distribution of selected PFAS and BAs in CD progressors and HCs. **A-C**) Levels (log^2^ intensities) of selected PFAS in cord blood (0 Mo) and at 3 months (3 Mo) of age in the CD progressors (CD) and HCs. **D-F)** Levels (log^2^ intensities) of selected BAs in the cord blood and at 3 months of age. The red dotted line denotes the mean of the population. The black solid lines in the bean plots represent the group mean. The group mean difference between case-control at a particular time-point was tested by unpaired two-sample t test. Mean difference between cord blood and 3 months samples obtain from the same infants were tested by paired t test. CD, HCs and BA denotes CD progressors, healthy controls and bile acids respectively.

In addition to the two primary BAs, we measured five primary/conjugated and secondary BAs (**Supplementary Table S2**). The levels of secondary BAs were mostly below the detection limits, particularly in the cord blood samples, where the total BA pool consisted mainly of GCA and GCDCA. Similarly, as for PFAS, no significant differences were observed between cases and controls either at birth or at 3 months of age (**Figure 1D-F**). The BA profiles at birth and at 3 months of age were markedly different in both groups (**Figure 1D-F**). The total BA pool showed significant, almost sevenfold upregulation (p = 0.0001). Particularly, GHCA and CDCA, which were close the limits of detection at birth, have increased substantially with age.

### Associations between PFAS, BA and lipid levels

Partial correlation network analysis between PFAS, BAs and lipids (classes) measured incord blood showed a different pattern of association in HCs (**Figure 2A)** and CD progressors (**Figure 2B)**. In the HCs, the level of deoxycholic acid (DCA) was inversely asociated with the sex of the infant, whereas no such association was seen in the CD progressors. Interestingly, DCA concentration was inversely associated with the level of PFOA. The CD group showed a strong association between age at diagnosis and PFOA exposure at both time points (**Figure 2B, Supplementary Figure 1**). In general, there was direct association between PFAS and several lipid classes. Age at diagnosis was also inversely associated with total TGs at birth. The association showed a clearly different pattern in the controls, both at birth and at the later time point (**Figure 2**).

**Figure 2.**
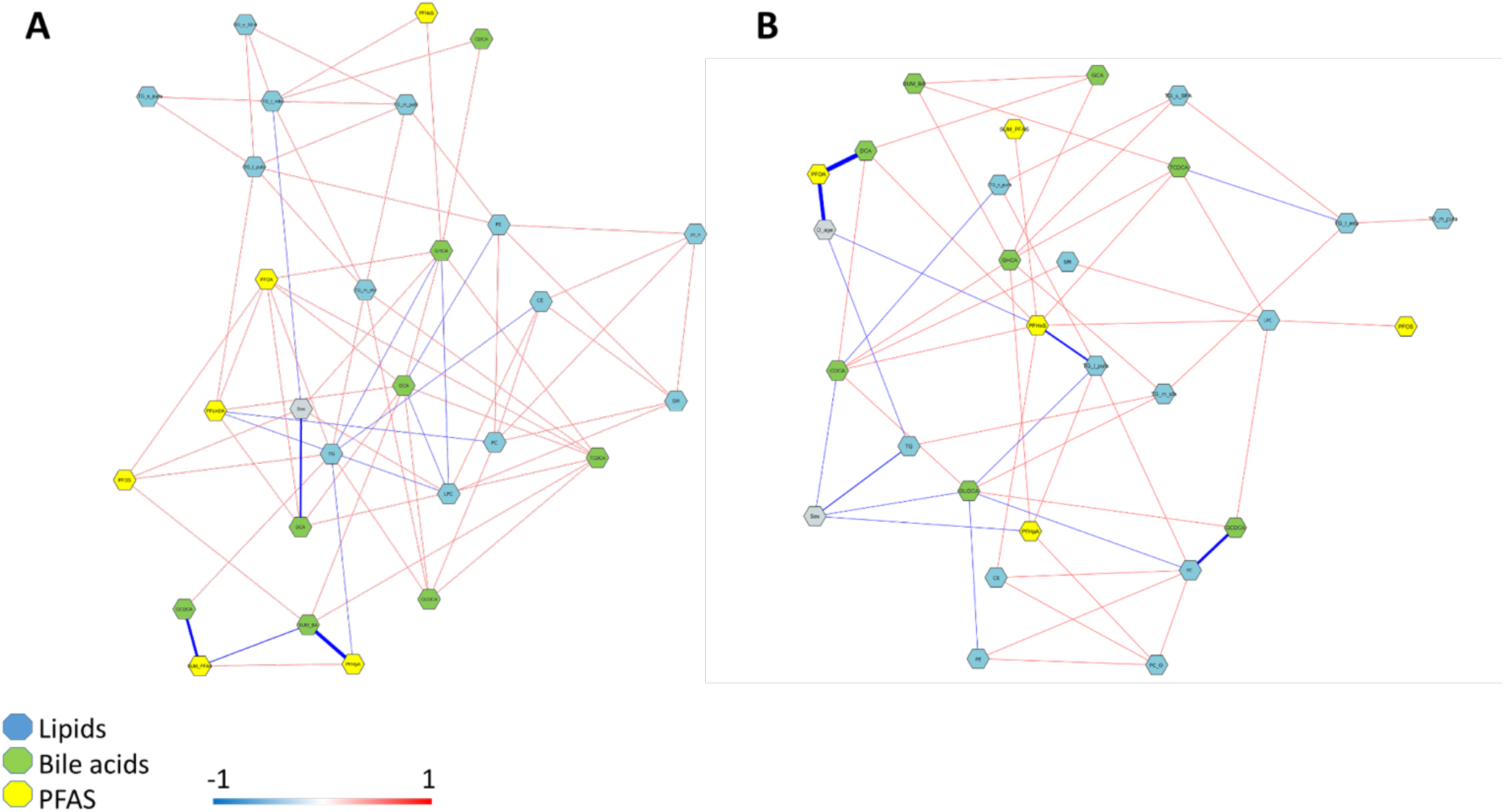
Molecular network of PFAS (yellow color), BAs (green color), lipid classes (grey color), and clinical variables (orange color) measured in the cord blood: **A)** Healthy controls. **B)** CD progressors. Red and blue lines denote positive and negative correlations respectively. The width of the lines represent the strength of correlation.

To understand how the individual PFAS and BAs are associated with the molecular lipid species, we performed multivariate^33^ and bivariate correlation analysis. Bivariate correlation analysis identified different correlation patterns in HCs and in CD progressors **(Figure 3**). In the control group there were significant associations mainly between the levels of the lipid classes at birth, and only the total concentrations of TGs showed associations with BAs (**Figure 3A**). In the CD progressor group, we observed significant correlations between the levels of BAs (GUDCA, GHCA, CDCA) and with multiple lipid classes in cord blood (PE, TG, SM) (**Figure 3B**). At 3 months of age, the patterns were different from those seen at birth. In the control group, specific BAs (CDCA, GUDCA, TCDCA and GCHA) showed a significant association with lipid levels, mainly with cholesterol esters (CEs) and TGs, with variable patterns depending on the fatty acyl saturation level (**Figure 3C**). Only a weak correlation was observed between the levels of PFAS and LPCs, while overall the PFAS levels were not associated with either lipids or BAs. In CD progressors, the patterns were different from the controls also at 3 months of age (**Figure 3D**). In this group, PFAS (PFOS and PFUnDA) levels and concentrations of TGs showed significant correlations, particularly with TGs containing saturated FAs. In addition, concentrations of BAs were also associated with those of TGs (DCA, GCA, CDCA and TCDA, total BAs), the directions being dependent on the bile acid species.

**Figure 3.**
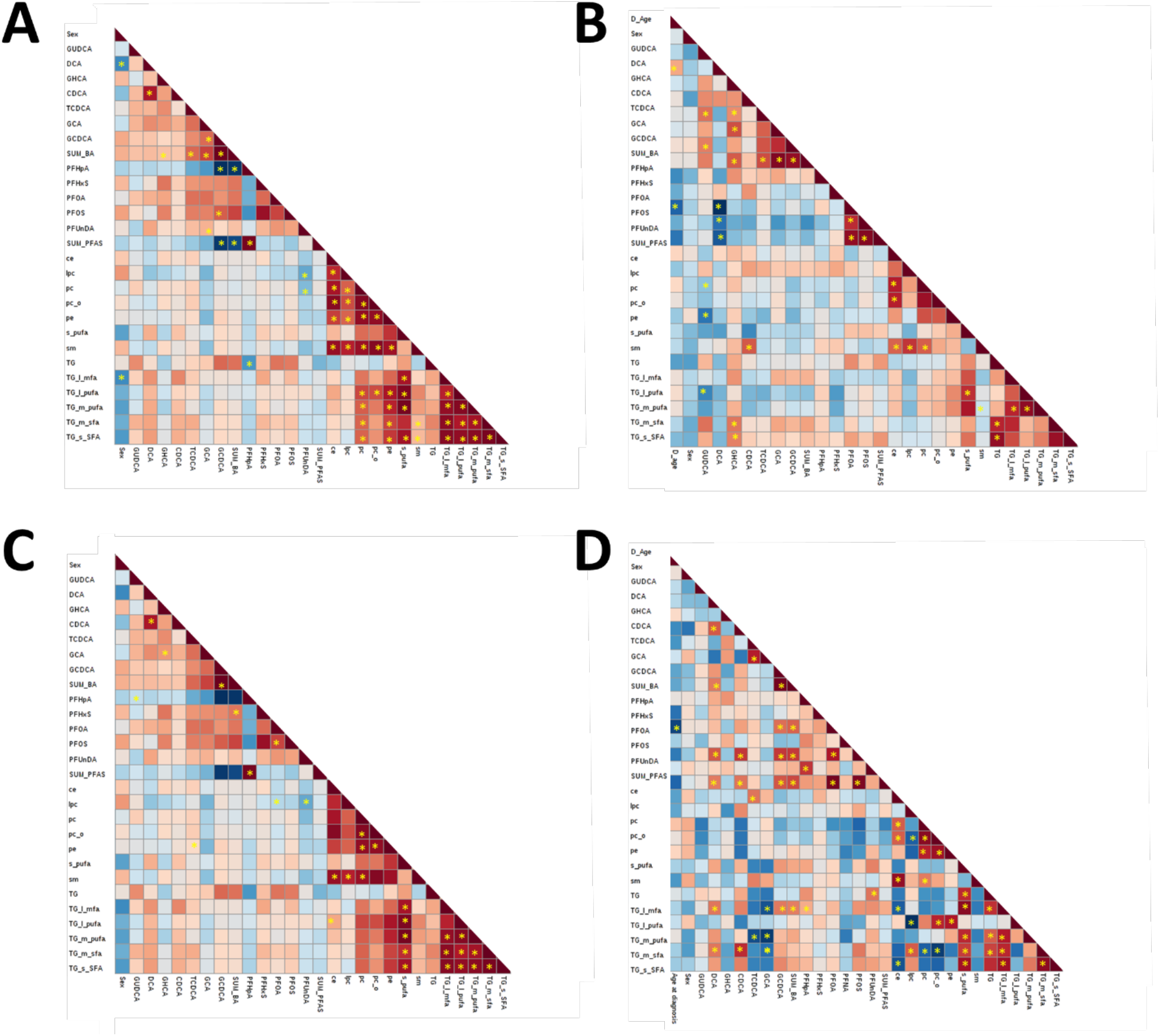
Correlation plots of PFAS, BA and lipid classes in (A) Controls at birth, (B) CD at birth, (C) controls at 3 months and (D) CD at three months. Red and blue colors denote positive and negative correlations respectively. Significant Pearson correlations are marked with *.

The main difference between the groups was the absence of correlation between the LPCs and the other phospholipid classes (PC, SM, PC) in CD progressors, while there was a strong and significant co-regulation of these lipid classes in the control group. At birth, there were no significantly different individual lipids between the groups^5^, however, the LPCs showed overall increased levels and most of the PCs decreased levels. LPC/PC ratio, with selected PC species containing PUFAs, showed significant upregulation in the CD group (FC=1.3, p=0.01).

### Molecular Predictors of PFAS, BA and Lipids in CD progression

Multivariate correlation analysis showed strong associations (r > 0.8) among PFAS, BAs and molecular lipids such as cholesterol esters (CEs), lysophosphatidylcholines (LPCs), phosphatidylcholines (PCs), phosphatidylethanolamines (PEs), sphingomyelin (SMs) and triacylglycerols (TGs) **(Figure 4A**). These associations become more complex at 3 months of age **(Figure 4D**).

**Figure 4.**
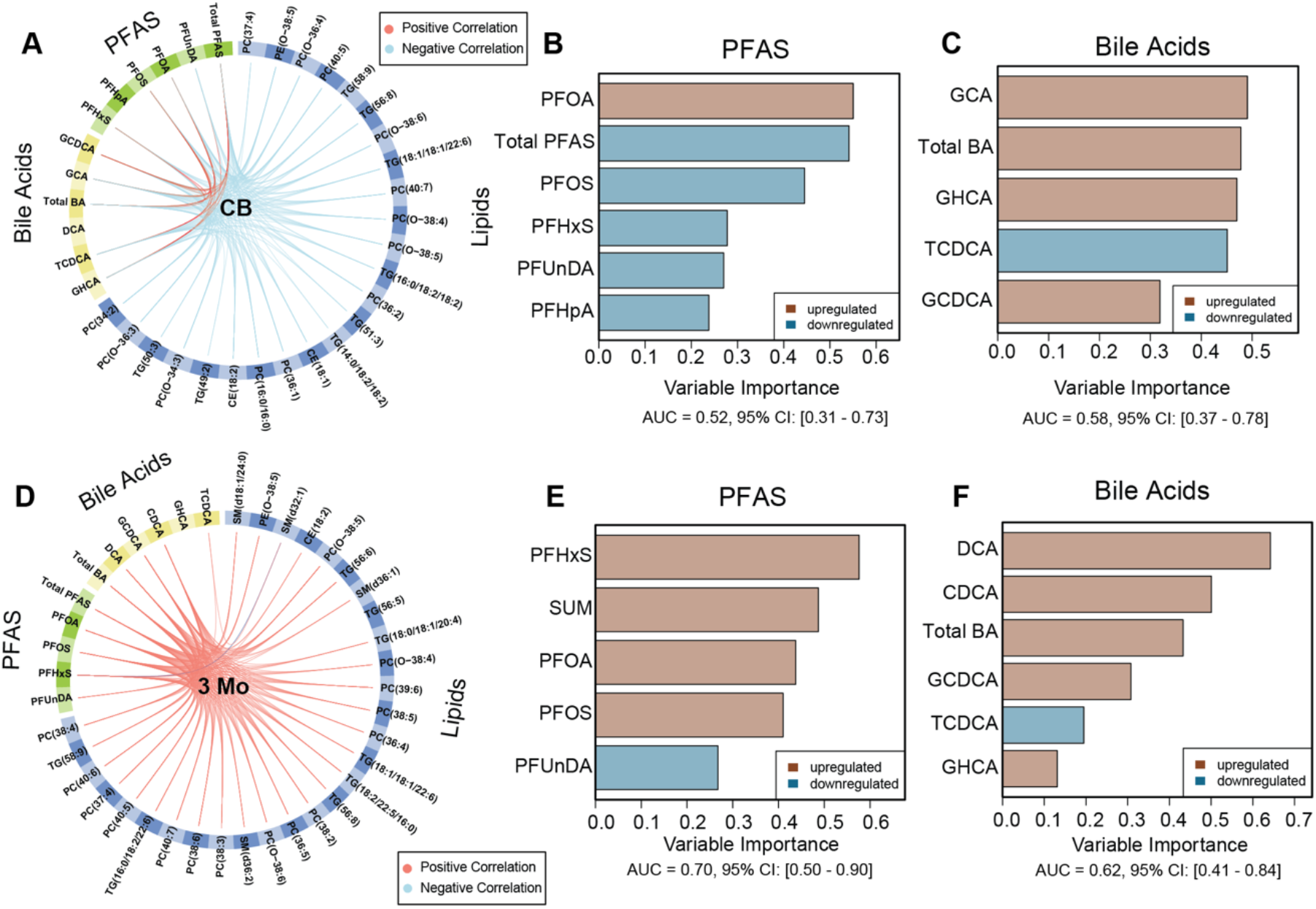
Integrative multiblock component analysis of PFAS, BAs and molecular lipids in the CD progressors *vs*. HCs. **A, D**) Circos plots (2 principal components) showing overall association (full-design matrix) between covariates of PFAS, BAs and molecular lipids in cord blood(CB), and at 3 months of age. Red and blue line denotes positive and negative correlations respectively. Sparse generalized correlation^33^ (coefficient, r > 0.5) are shown. **B-C)** Horizontal barplots showing top selected predictors/contributors/discriminators from each PFAS and BA data blocks, where the length of bar corresponds to the absolute loading weights (variable importance) of the feature, that tends to discriminate CD progressors *vs*. HCs at birth. The direction of regulation of the predictors are shown with light brown and blue colors that corresponds to the CD progressors or HCs respectively; wherever the mean intensity of the predictor is maximum. **E-F)** Key predictors of PFAS and BAs, that helped to classify CD progressors vs. HCs at 3 months (3 Mo) of age. The model performance on each block of data are given by cumulative (principal components) area under the curves (AUCs).

Next, we selected those lipids that showed significant differences between CD and HCs in infancy in our previously reported study^5^. These lipids together with the BAs and PFAS were subjected to multiblock (MB) analysis. MB analysis identified PFAS, BA and molecular lipid species as predictors (discriminative features) that helped to classify CD progression *vs*. HCs. The association between top predictors (high variable importance scores) are shown (**Figure 4A, D**).

In cord blood, the levels of PFOA and PFOS were identified as the top linear predictors of CD progression (**Figure 4B)**. On the other hand, among the BAs, the levels of primary BAs such as GCA and GHCA are the strongest predictors of CD progression (**Figure 3C)**. Moreover, elevated concentration of PFOA in the CD progressors was positively associated with the levels of GCA and GHCA. This suggests that prenatal exposure to PFOA and/or PFOS might accelerate the risk of CD progression. Presumably, it alters the BA and lipid metabolism. Interestingly, lipids such as CE(18:1) (intermediate of BA synthesis) were identified as predictors of CD progression; the levels of PFOA and PFOS were inversely correlated with those of CEs (**Figures 4A** and **5A**). In addition, many ether-linked PCs such as PC(O-34:3), PC(O-36:3,4) and PC(O-38:4,5,6) were identified as key contributors, that were downregulated in the CD progressors as compared to the HCs (**Figures 4A** and **5A**).

At 3 months of age, concentrations of PFHxS was marked as a top predictor associated with CD progression, along with levels of PFOA and PFOS. Moreover, the level of PFHxS were increase in the CD progressors as compared to HCs (**Figures 1A, 4E** and **5B**). At this age, elevated levels of secondary BAs such as DCA and CDCA were marked as the key predictors for CD progression (**Figure 4F**). As stated before, the level of PFOA was inversely correlated with that of DCA **(Figure 2B)**, suggesting an interaction between the PFAS and BAs in the CD progressors. Moreover, PFOA was inversely linked to age at diagnosis of CD (**Figure 2B** and **Figure 6**). In addition, the MB model identified several PCs, SM(d36:2), PC(O-38:4,5) as key predictors that discriminate the lipid profiles of CD progressors from those seen in the HCs (**Figure 5B**).

**Figure 5.**
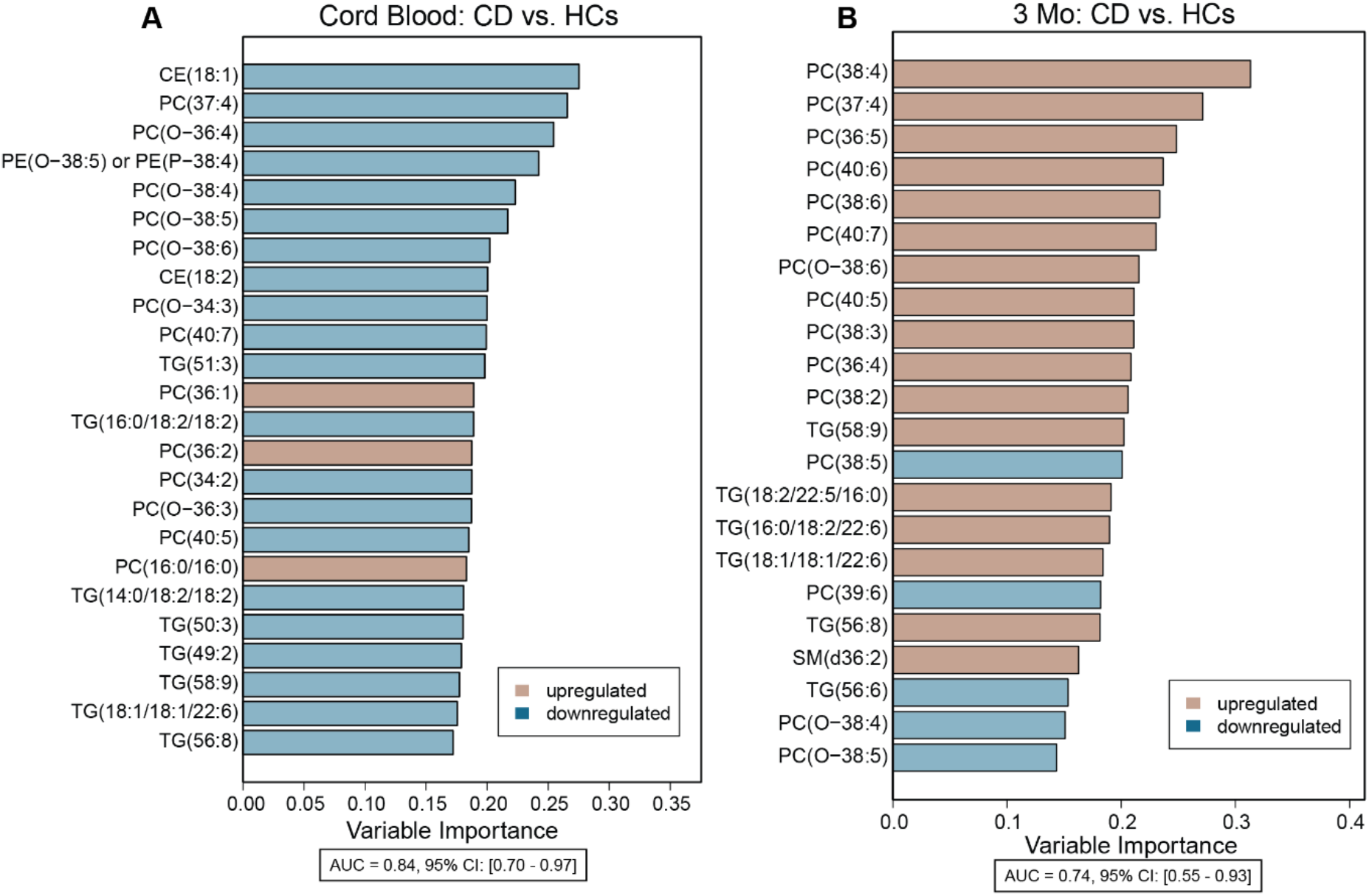
Key lipid contributors/predictors, that jointly (together with PFAS and BAs), aided in the separation of CD progressors *vs*. HCs, at birth and at 3 months of age. The direction of regulation of the predictors are shown with light brown and blue colors that corresponds to the CD progressors or HCs respectively; wherever the mean intensity of the predictor is maximum.

**Figure 6.**
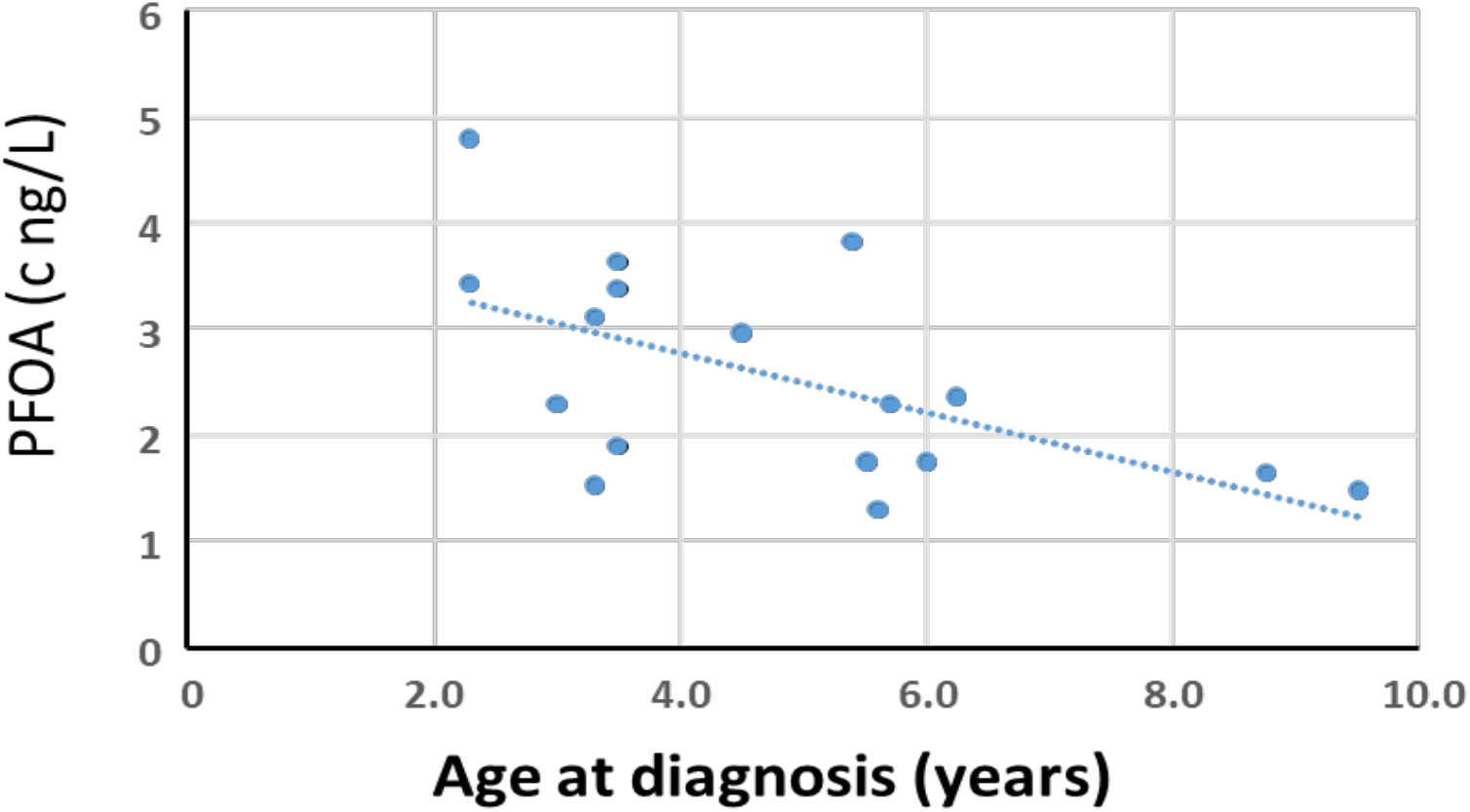
Correlation of PFOA concentration at birth and the age of diagnosis. Pearson correlation r=-0.575.

## Discussion

We observed dysregulation between intralipid class regulation at birth in those infants that progressed to CD, particularly in LPC/PC ratios. PFOS and PFHxS levels were also positively associated with LPCs in the CD progressors, but not in HCs. However, 3 months later, the ratio was similar in both groups. Increased LPCs have been indicated to be characteristic for CD progression at later time points, both our earlier study as well as in another recent study showed increased LPCs already in those 4-month-old infants who progressed later to CD^21^. Furthermore, our results implied that the associations between BAs and complex lipids show different patterns already at birth in those infants who later developed CD as compared with healthy controls. Significant correlations were observed between TGs and specific BAs already at birth in the CD progressors. At 3 months of age, both groups showed a significant correlation between the lipids and BAs, although the patterns were different. At this later time point, the CD group showed significant positive correlations between different classes of TGs while the healthy controls showed inverse correlations. Thus, the data agrees well with our earlier study, in which we observed distinct lipid changes, particularly in the TG class, in children who later progressed to CD already prior to the first exposure to dietary gluten. The data suggested that the specific TGs, found elevated in the CD progressors, may be due to a host response to compromised intake of essential lipids in the small intestine, requiring *de novo* lipogenesis. The current study suggests that the changes in TG metabolism are also related to alteration in BA metabolism, and that early-life exposure to PFAS may contribute to these changes. Currently, there is very limited amount of data on BA and lipid metabolism in infants. The BA pool in the fetus and in newborn infants is unique. More than 90% of fetal BAs represent conjugated forms of the primary BAs, cholic acid (CA) and chenodeoxycholic acid (CDCA), since the intestinal bacteria necessary to transform primary BAs into secondary BAs are thought to be absent *in utero*, Based on recent findings, the fetal BAs may be very different than later in life^37,38^. The enterohepatic circulation of BAs *in utero* is also minimal. Instead, there is a transplacental gradient for BAs in the fetal-to-mother direction, except for secondary and tertiary BAs, which are more abundant in maternal serum^39^.

PFAS exposure has been shown to impact BA metabolism through several mechanisms. BAs and PFAS have been shown to have a similar enterohepatic circulation^25,28^, and it has been indicated that 7-alpha-hydroxylase (CYP7A1), which catalyzes the first and rate-limiting step in the classical pathway in the formation of BAs from cholesterol, may be down-regulated by PFAS^25,40^. This might lead to increased re-uptake of BAs, which would generate negative feedback loops *via* the farnesyl-X-receptor and subsequently reduce their *de novo* synthesis^25,41^. It has also been shown that PFOA inhibits the function of the hepatocyte nuclear factor 4α^42^, which plays a central role in the regulation of BA metabolism in the liver, and is linked both with the synthesis and conjugation of primary BAs. However, in neonates, the alternative (acidic) pathway is the major pathway for bile acid synthesis and this pathway is governed by mitochondrial sterol 27-hydroxylase (CYP27A1), which can initiate a process independent of CYP7A1. Only after weaning, CYP7A1 is expressed and the classic pathway becomes the major pathway for bile acid synthesis in the liver^25^. There is currently no data on the impact of PFAS exposure on the regulation of the CYP27A1. On the other hand, the impact of PFAS on the TG metabolism may be modulated through the bile acids, as the bile acid receptor FXR has also a regulatory role in triglyceride metabolism^43-45^. In humans, recent data suggests that FXR is not activated directly by PFAS^43^. In the liver, FXR activation, by, *e*.*g*., bile acids, would result in downregulation of CYP7A1, which in addition to inhibition of the classical BA synthetic pathway also reduces the expression of several genes mediating free fatty acid synthesis, thereby attenuating *de novo* lipogenesis^25^. Thus, activation of FXR modulates free FA oxidation and the clearance of TG to the circulation.

The acylalkyl PCs type of lipids that showed significant association with the CD progression were positively associated with the levels of total BAs and PFHxS, indicating potentially disturbed hepatic synthesis of these lipids, particularly as the intra-lipid regulation between these lipids and particularly TGs was disturbed in CD progressors. These ether lipids are perceived to function as endogenous antioxidants in addition to their structural roles in cell membranes, and emerging studies suggest that they are involved in cell differentiation and signaling pathways as well^46^. Interestingly, these lipids have been observed to be endogenous antigens to activate invariant natural killer T cells (iNKT)^47^, which are subset of innate immune cells. Recently, therapeutic potential of iNKT cell antigens against autoimmunity have been implicated^48^. Thus, the reduced levels of the alkyl ether lipids could suggest a compromised response to oxidative stress. Interestingly, these lipids have previously been found to be associated with disease development also in other autoimmune diseases, such as type 1 diabetes^49^, thus indicating that they may play an important role in the development of autoimmune disorders. Due to their role in cell membranes, the decrease in ether-linked PCs particularly, PC(O-38:4,5) might also attribute to compromised intestinal permeability in the CD progressors.

Taken together, our results show that PFAS exposure may modulate lipid and BA metabolism, and the impact is different in the infants who develop CD later in life, when compared with healthy controls. Although we did not observe any significant differences in the levels of PFAS exposure in those children that later developed CD, which may be due to the small sample size, we did observe a significant increase in their levels in progressors to CD from the time of birth to 3 months of age, suggesting more efficient uptake of PFAS. Furthermore, age at diagnosis of CD was strongly associated with the PFAS exposure. Our study thus suggests that further investigations of the impact of exposures to environmental chemicals in the development of autoimmune diseases are merited.

## Data Availability

The lipidomics data sets and the clinical metadata generated in this study are available from MetaboLights, under accession number MTBLS729.

## Conflicts of interest

### Guarantor of the article

Tuulia Hyötyläinen, Ph.D.

### Specific author contributions

T.H., M.K., M.O.: conception and design of the study; L.S., S.S., S.V., H.H., J.I., J.T., R.V., M.O., M.K., T.H.: generation, collection, assembly, analysis, and/or interpretation of data; T.H. and M.O.: drafting or revision of the manuscript; all authors: approval of the final version of the manuscript.

### Financial support

This study was supported by funding from VetenskapsrÅdet (to T.H.; grant no. 2016-05176) and Formas (to T.H. and M.O.; grant no. 2019-00869). The DIPP study was supported by the following grants: JDRF (grants 1-SRA-2016-342-M-R, 1SRA-2019-732-M-B); European Union (grant BMH4-CT98-3314); Novo Nordisk Foundation; Academy of Finland (Decision No 292538 and Centre of Excellence in Molecular Systems Immunology and Physiology Research 2012-2017, Decision No. 250114) and Special Research Funds for University Hospitals in Finland.

## Acknowledgments

We thank the families participating in the DIPP study for making this study possible. We also thank the expert staff of the DIPP study for their excellent work with the participating research families and sample collection.

---

**Supplementary Figure 1.**
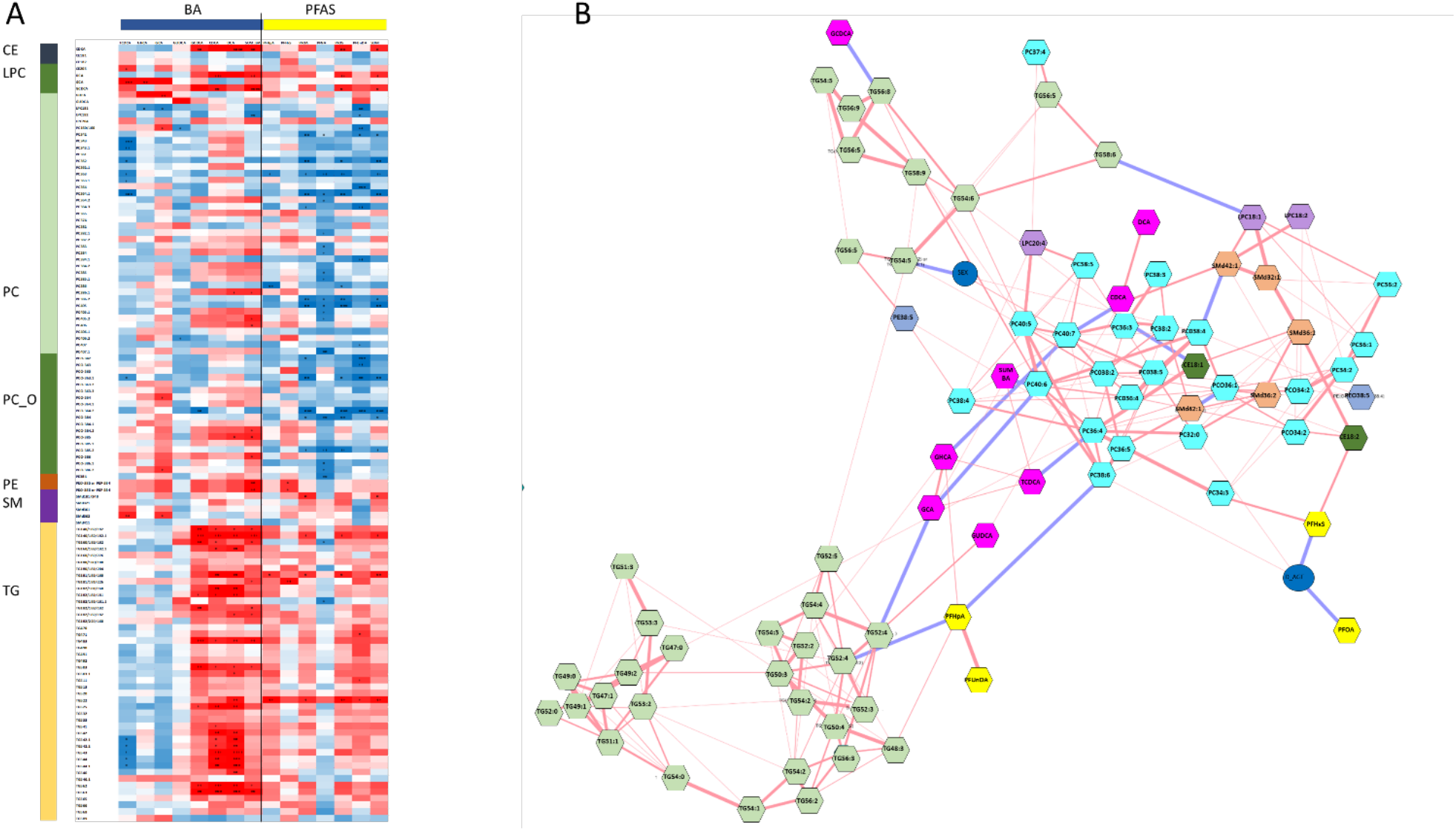
(A) correlation plots of lipid-BA and lipid-PFAS (3 months), Pearson correlations, significant correlations marked with * (p<0.0001 ****, p<0.001 ***, p<0.05 **, p<0.1*) and (B) Partial correlation network of the previously identified panel of CD-related lipids, BAs and PFAS (at birth and at 3 months of age). Significant correlation was observed in the CD group between BAs and the lipids. Particularly, GCDCA, CDCA and DCA showed significant positive correlations with TGs of low carbon number and double bond count, while TCDCA and GUDCA showed inverse associations with PCs at 3 months of age. At the same time point, PFOA, PFOS and PFUnDA, and total PFAS showed positive correlations with the TGs containing low carbon number and double bond count and inverse associations with PCs. In the partial correlation network, PFOA and PFHxS showed an inverse association with the age at diagnosis and PFHxS further positive associations with CE(18:2) and PC(34:3) (**Figure 4B**). PFHpA showed an inverse association with one TG and one PC species and a positive association with GHCA. BAs showed mainly inverse associations with several lipid species.

**Supplementary Table S1.**
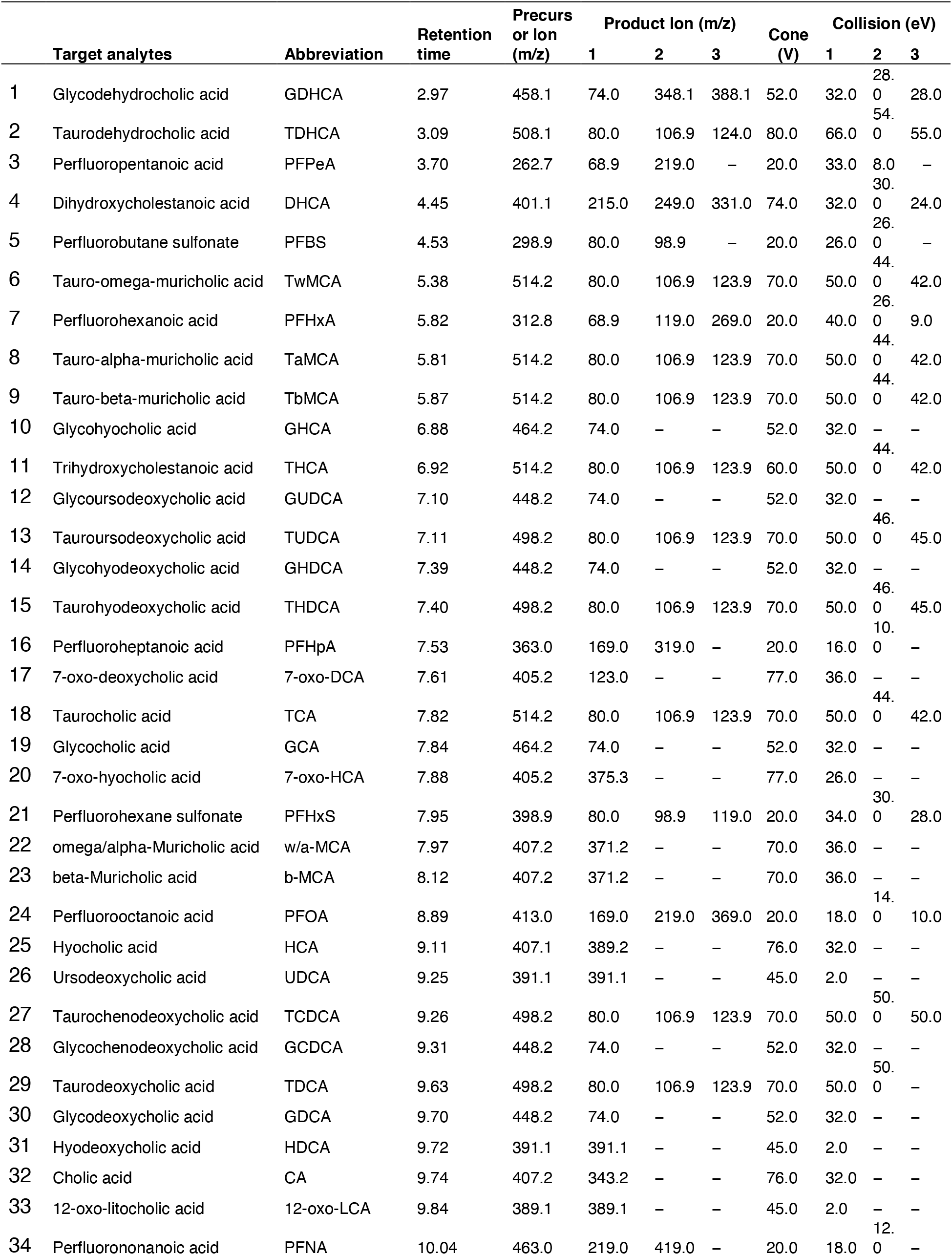
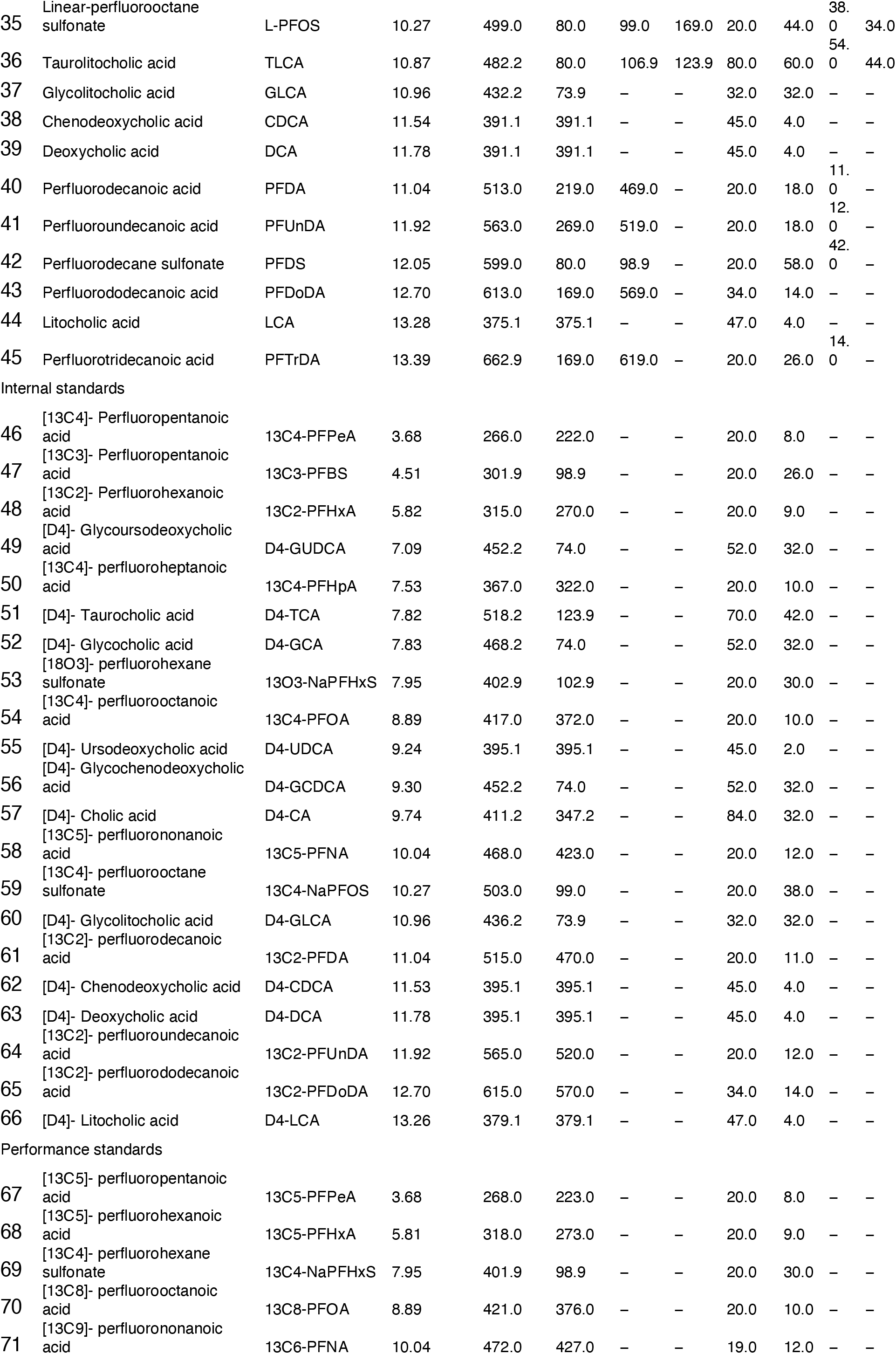
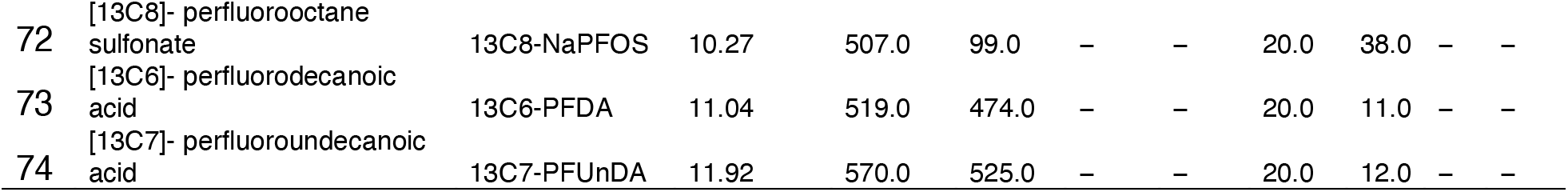
Acquisition parameters including the list or targets compounds ordered by retention time.

**Supplementary Table S2.** PFAS concentrations.

|  | CTRL_CB |  |  | CD_CB |  |  | CTRL 3 months |  |  | CD 3 months |  |  |
| --- | --- | --- | --- | --- | --- | --- | --- | --- | --- | --- | --- | --- |
|  | median | min | max | median | min | max | median | min | max | median | min | max |
| **PFHpA** | <LLQ | <LLQ | 378.39 | <LLQ | <LLQ | 0.5 | <LLQ | <LLQ | 0.69 | <LLQ | <LLQ | 1.09 |
| **PFHxS** | 0.49 | 0.31 | 0.99 | 0.55 | 0.31 | 1.03 | 0.59 | 0.31 | 0.93 | 0.70 | 0.31 | 1.62 |
| **PFOA** | 2.43 | 1.23 | 4.46 | 2.32 | 1.31 | 4.80 | 4.05 | 0.98 | 6.25 | 4.34 | 1.23 | 9.17 |
| **PFOS** | 2.25 | 0.27 | 5.32 | 2.21 | 0.27 | 8.17 | 3.40 | 0.71 | 6.70 | 2.93 | 0.27 | 7.66 |
| **PFUnDA** | <LLQ | <LLQ | 0.44 | <LLQ | <LLQ | 0.48 | <LLQ | <LLQ | 0.63 | <LLQ | <LLQ | 0.53 |
| **PFAS** | 6.04 | 3.19 | 11.69 | 6.87 | 3.42 | 14.17 | 9.74 | 3.82 | 12.77 | 9.15 | 3.17 | 18.55 |
| TCDCA | 32.42 | <LLQ | 32.42 | 17.30 | <LLQ | 17.30 | 44.01 | <LLQ | 44.01 | 72.22 | <LLQ | 72.22 |
| GHCA | <LLQ | <LLQ | <LLQ | <LLQ | <LLQ | <LLQ | 30.78 | <LLQ | 30.78 | 28.88 | <LLQ | 28.88 |
| GCA | 171.02 | <LLQ | 171.02 | 150.22 | <LLQ | 150.22 | 88.62 | <LLQ | 88.62 | 252.44 | <LLQ | 252.44 |
| GUDCA | <LLQ | <LLQ | <LLQ | <LLQ | <LLQ | <LLQ | 0.65 | <LLQ | 0.65 | 1.58 | <LLQ | 1.58 |
| GCDCA | 205.30 | <LLQ | 205.30 | 135.63 | 16.87 | 135.63 | 862.43 | 138.38 | 862.43 | 1391.33 | 176.30 | 1391.33 |
| CDCA | <LLQ | <LLQ | <LLQ | <LLQ | <LLQ | <LLQ | 108.38 | <LLQ | 108.38 | 126.32 | <LLQ | 126.32 |
| DCA | <LLQ | <LLQ | <LLQ | <LLQ | <LLQ | <LLQ | <LLQ | <LLQ | <LLQ | 3.58 | <LLQ | 3.58 |
| SUM BA | 465.28 | <LLQ | 465.28 | 392.87 | 16.87 | 392.87 | 1210.23 | 313.41 | 1210.23 | 2267.03 | 403.53 | 2267.03 |

**Supplementary Table S3.**
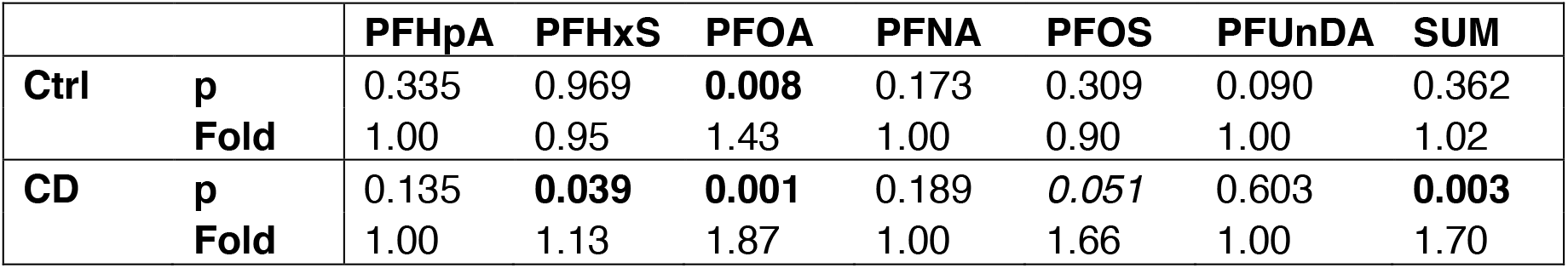
Change of PFAS levels from cord blood to 3 months.

